# Explainable Transformer-Based Neural Network for the Prediction of Survival Outcomes in Non-Small Cell Lung Cancer (NSCLC)

**DOI:** 10.1101/2021.10.11.21264761

**Authors:** Elly Kipkogei, Gustavo Alonso Arango Argoty, Ioannis Kagiampakis, Arijit Patra, Etai Jacob

## Abstract

In this paper, we introduce the “Clinical Transformer” - a recasting of the widely used transformer architecture as a method for precision medicine to model relations between molecular and clinical measurements, and the survival of cancer patients. Although the emergence of immunotherapy offers a new hope for cancer patients with dramatic and durable responses having been reported, only a subset of patients demonstrate benefit. Such treatments do not directly target the tumor but recruit the patient’s immune system to fight the disease. Therefore, the response to therapy is more complicated to understand as it is affected by the patient’s physical condition, immune system fitness and the tumor. As in text, where the semantics of a word is dependent on the context of the sentence it belongs to, in immunotherapy a biomarker may have limited meaning if measured independent of other clinical or molecular features. Hence, we hypothesize that the transformer-inspired model may potentially enable effective modelling of the semantics of different biomarkers with respect to patients’ survival time. Herein, we demonstrate that this approach can offer an attractive alternative to the survival models utilized in current practices as follows: (1) We formulate an embedding strategy applied to molecular and clinical data obtained from the patients. (2) We propose a customized objective function to predict patient survival. (3) We show the applicability of our proposed method to bioinformatics and precision medicine. Applying the clinical transformer to several immuno-oncology clinical studies, we demonstrate how the clinical transformer outperforms other linear and non-linear methods used in current practice for survival prediction. We also show that when initializing the weights of a domain-specific transformer by the weights of a cross-domain transformer, we further improve the predictions. Lastly, we show how the attention mechanism successfully captures some of the known biology behind these therapies.

## 1 Introduction

In recent years, computational modelling of sequences with variable lengths and long-range dependencies received a boost through the introduction of a new class of architectures called transformers. These models were effective in contextual modelling on account of their self-attention architecture that allowed them to differentially focus on relevant parts of an input sequence. Such an ability to utilize relevant parts of an input, where such salient sub-sections may not be sequentially located, enables the transformer to approach the modelling of variable length inputs using the semantic context implied therein.

Motivated by the applicability of self-attention, we extend to the domain of precision medicine and ask whether it is possible to predict treatment outcome and response from individualized records of patients undergoing immuno-oncology interventions. Such records aggregate disparate information about patient clinical assessments, demographics, treatment history, along with various tumor molecular measurements and can be combined in a sequence and formulated as a machine learning problem with a response variable such as overall survival (continuous) or response (categorized or binary). Importantly, at the single patient level, these clinical and molecular features conceal complex interrelationships that are intertwined with the patient outcome. The question then distils down to the choice of a suitable learning machine that can approximate such undetermined interactions across the space of the available clinical and molecular data for individual subjects. This is achieved in our case through a novel interpretation of the transformer architecture proposed in this work.

A frequent problem in clinical studies, especially in early stage (that is, phase I and II) trials, is the small sample size which poses a great challenge to building reliable and interpretable prediction models. In addition, the complexity of the underlying biology and the heterogeneity of the patient population impair the ability to define biomarkers that can be used for patient selection in subsequent trials. Therefore, the ability to utilize transfer learning, an attribute that comes as part of the transformer architecture, is of great importance, especially since the majority on the cancer drugs, including immuno-oncology compounds are only effective in a small subset of the patients. Importantly, transferable representations enable us to extend automated survival analysis to conditions where limited patient data is available in a manner that was not possible in traditional approaches. This also allows future extensions of our precision medicine approach for orphan or rare diseases where the personalization of care is often also hampered by limited data availability.

## 2 Related Work

The ability of transformer architectures to handle sequences without requiring intermediate storage represents a significant shift in how machine learning models process such sequences, as is often a requirement in several applications in systems biology. Transformer architectures introduced a significant acceleration in the development of computational models for holistic understanding of sequential data. Originally proposed for NLP by Vaswani et al. [2017], transformers have been extensively applied to other fields (Chaudhari et al. [2019]). Furthermore, a number of studies have utilized transformers or a modified version to address biological problems. For example, Widrich et al. [2020] implement a modern Hopfield Network with transformer-like attention for immune repertoire classification. Elnaggar et al. [2020] trained Trasformer XL (Dai et al. [2019]), BERT (Devlin et al. [2018]) and ALBERT (Lan et al. [2020]) on protein sequences to predict secondary structures; Avsec et al. [2021] implemented enhancer transformer to predict gene expression from DNA sequence; Jumper et al. [2020] predicted protein structure using 3D equivariant transformers.

Survival analysis is an area that has been dominated by statistical approaches (Harrell Jr [2015], Cox [1972]) such as multi-variate modelling using Cox proportional hazards (CoxPH) for predicting patients’ response to therapy based on individual’s genomic and clinical features (Anagnostou et al. [2020]). Several non-linear approaches have also been proposed including gradient boosting machines (Pölsterl [2020]) among others. In recent years, deep neural networks have also been applied to survival analysis. For instance, Katzman et al. [2018], developed DeepSurv, a personalized treatment recommender system using a CoxPH deep network. Yousefi et al. [2017] implemented SurvivalNet that uses Cox partial-likelihood to train a neural network to predict survival outcomes. Hu et al. [2021] developed a transformer-based survival model that utilizes ordinal regression to optimize survival probabilities over time. However, none of these methods take into account the interactions between the features in an explicit way as part of the model or enable reviewing of the model at the single patient level.

With the objective of personalized risk prediction in mind, we propose our key contributions as follows:

- A novel approach based on transformer architectures that can handle both clinical and molecular features and incorporate a custom loss function that links together the input features, internal attention mechanisms and clinical endpoints (time to a clinical event).
- A framework that explains and summarizes the mutual relationships between different features and the relationships between features and clinical endpoints, as opposed to the state-of-the-art competing frameworks that render themselves incompatible with downstream explainability as required in several precision medicine applications Maciejko et al. [2017].
- A widely applicable framework in computational biology that is validated on public data sets with demonstrated use cases in precision medicine and bioinformatics.

## 3 Methods

### 3.1 Training and Dataset

The general framework for clinical transformer training and testing is described in Figure 1. A large cohort of 1661 patients from the Memorial Sloan Kettering Cancer Center (MSKCC) comprising of advanced cancer patients treated with Immune Checkpoint inhibitor (Figure 1A) was downloaded for analysis (Samstein et al. [2019]). Each patient entry included genomic information of 468 genes (those are, mutation calls for each of the 468 genes) and 8 clinical features (e.g. age and sample type) (See further details in Samstein et al. [2019]. To reduce sparsity in the data, genes with mutations in less than 5% of the total population were excluded from further analysis, resulting in 55 features with genomic information instead of the initial 468. A cross-domain clinical transformer (Figure 1B) was pre-trained on 10 cancer types (N=1266) and a “snapshot” of NSCLC patients (50 randomly selected NSCLC patient out of a total of 344 patients). Later, specific clinical transformer (Figure 1C) was trained with 80% of the NSCLC data excluding the 50 patients that were used for the model in B. This transformer was initialized with the weights from the pan cancer transformer. Evaluation was performed by the concordance index over the testing set (20%) of the NSCLC clinical transformer B (Figure 1D). To account for stability, this process was repeated 100 times with randomly generated 80/20 splits. Transformer attentions capture interactions between input features in the context of patients’ survival (Figure 1E). These attention weights can be used for different bioinformatics applications, including network analysis in systems biology, gene set enrichment to identify dominant biological pathways with clinical relevance, clustering and other pattern recognition methods for patient stratification.

**Figure 1:**
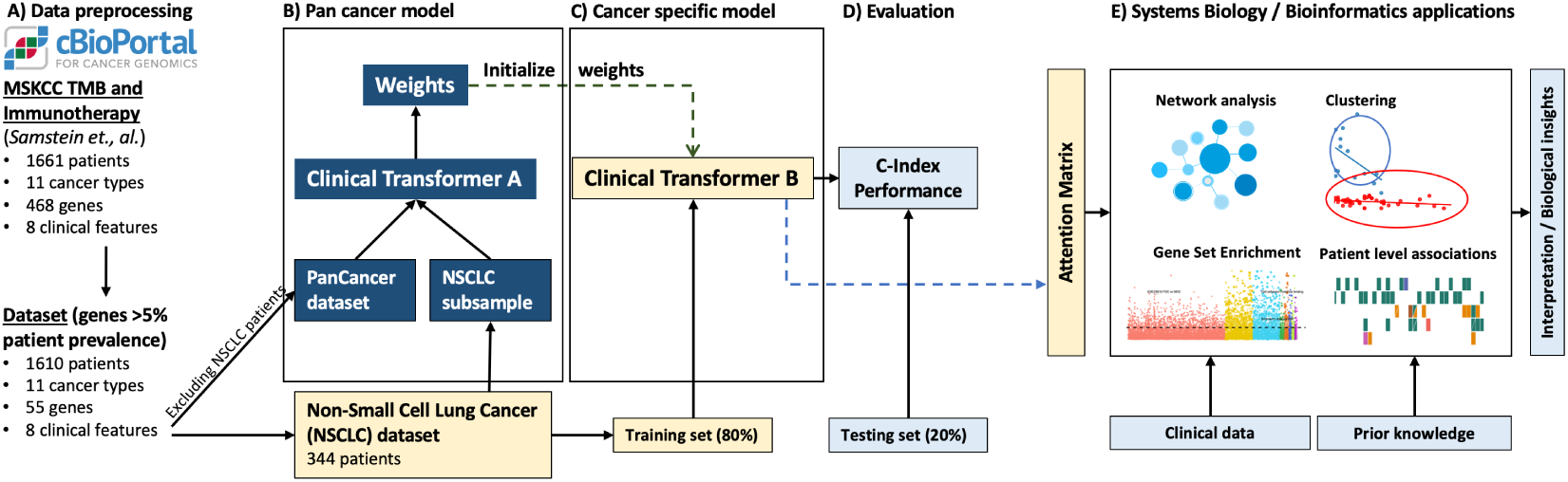
General framework for the Clinical Transformer. A) Data preprocessing. B) Training of pan-cancer model using a cross-domain clinical transformer. C) Training of cancer-specific model on 80% of the NSCLC. D) Evaluation of clinical transformer using concordance index. E) Applications of transformer attentions across different bioinformatics analysis.

### 3.2 Numerical embedding of the clinical and genomic feature space

The initial transformer architecture proposed by Vaswani et al. [2017] uses positional encoding vectors to account for the absolute location of tokens in the sequence. In our case, since we are not dealing with sequential data, we excluded the positional encoding vectors from the transformer architecture. To process numerical data, we projected the initial set of features *F* ∈ ℝ^*N ×*1^ (*F* is a vector of 65 features in our analysis) to an embedding space 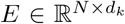 *E* is a matrix of 65 by 32 in our analysis) by feeding the feature space *F* into a dense layer (Figure 2A). Therefore, each component of the feature space *F* is decomposed into a linear combination of the learned weights that is then fed into the transformer. The rationale behind this decomposition is to embed numerical data into a fixed vector size.

**Figure 2:**
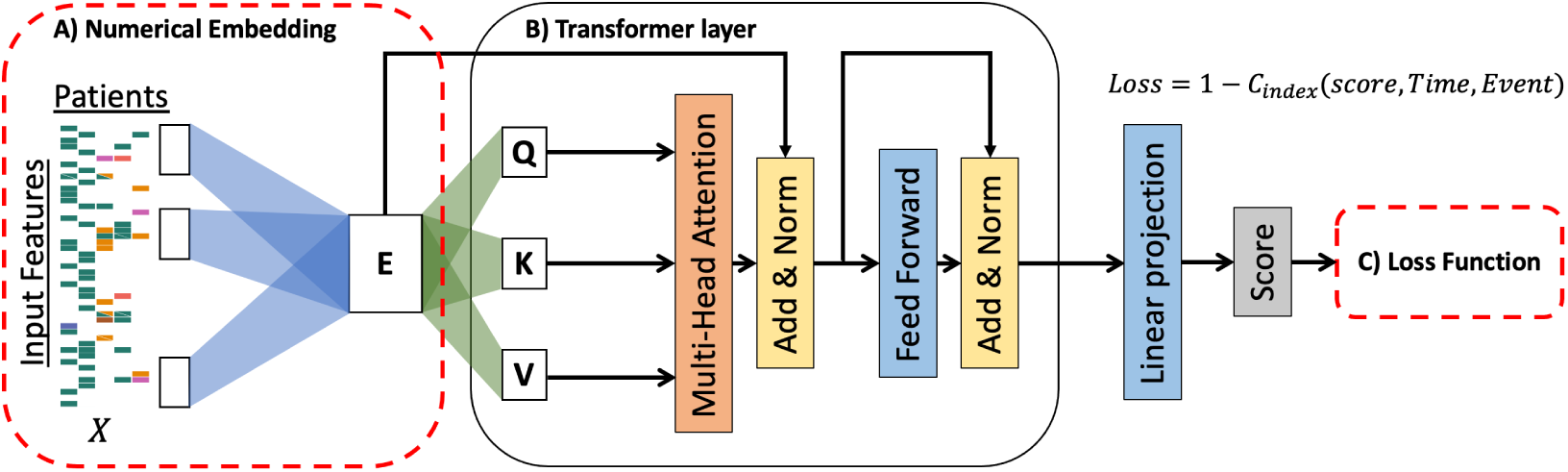
Schematic representation of the Clinical Transformer. The numerical embedding layer (A) takes tabular data as input and projects each feature into a d dimensional vector. The transformer layer (B) takes as input *N* by *D* matrix and projects it into the *K, Q* and *V* matrices that are fed into the transformer layer. Output of the transformer layer is flattened and translated into a risk score. The model is trained using the c-index-based loss function which takes into account survival time and censorship information.

### 3.3 Scaled Dot-Product Attention

The scaled dot-product attention in a transformer, enables the model to selectively focus on relevant features from the input space by identifying similarities among the input features while associating those similarities with the model outcome. The attention is defined as:

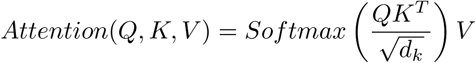

where *Q, K,* 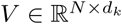. *N* is the number of input features and *d*_*k*_ is the dimension of key, query and value vectors (see Figure 2B).

### 3.4 Loss function

To optimize model parameters towards patient survival outcomes, instead of a binary response or text translation, we utilized the concordance metric in survival analysis workflow as a measure of model discrimination (Figure 2C). Harrell’s concordance index C is defined as the proportion of observations that the model can order correctly in terms of survival times (Steck et al. [2008]). This can be interpreted as a generalization of the area under the ROC curve (AUC) that considers censored data. It represents the global encapsulation of the model discrimination power and its ability to provide a reliable ranking of the survival times based on the individual risk scores. In our workflow, the concordance-based model discrimination was implementing using a loss function with a sigmoid approximation of Harrell’s C-index (Schmid et al. [2016], Mayr and Schmid [2014]). This led to an objective of the form:

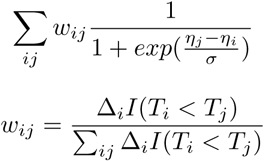

where the indices *i* and *j* refer to pairs of observations in the sample, Δ_*i*_ = 0 if censored and 1 if deceased, *T* is the corresponding survival time, *η* is the predicted scores from clinical transformer and *σ* is a smoothing parameter for the sigmoid approximation. *η* can be thought of as *xβ* where 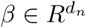 are the weights from the transformer’s last linear projection layer and *d*_*n*_ is the number of nodes.

The final outputs from clinical transformer are: (1) Predicted scores - output from the final layer of fully connected neural network with linear activation function. (2) A list of attention matrices of the form softmax 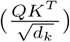. The length of this list corresponds to the number of transformer layers.

### 3.5 Hyperparameter tuning

We tested the clinical transformer using different settings, including different number of heads, layers, epochs and learning rates. We identified that the model quickly overfits after 200 epochs. We also observed that the model starts to overfit as the number of layers increase. We did not observe a significant difference in performance when using 4 or more heads per layer (see Figure S7). The optimal setting of the clinical transformer was set to 2 layers, 4 heads, 100 epochs and 0.001 learning rate. We trained our models in one machine with one Tesla 32GB V100 GPU.

## 4 Results and Discussion

### 4.1 The clinical transformer outperforms current practice of survival analysis in the field

The predicted scores from the transformer are directly proportional to patients’ survival time and therefore the scores can be analyzed using standard survival analysis approaches such as the concordance index (note that a concordance index of 0.5 indicates random predictions) Mayr and Schmid [2014]. Alternatively, we can stratify patients into low and high scores using the predicted scores while maximizing the separation of the Kaplan Meier (KM) curves between these groups. Here, we utilized these two approaches to assess the performance of the predictions. We computed the concordance index (CI) for every test set prediction in the 100 training-test random splits (referred to as “folds” in the rest of the paper). We also performed patient stratification using the test sets predicted scores and computed corresponding hazard ratios comparing high vs low scores. To test model’s consistency in assigning scores to different patients, we tracked each patient in the test set across the folds. These patients were then subdivided into four ‘survival’ groups: super-responders (*>* 18 months), two intermediate group (*≥* 12 months & *≤* 18 months, *≥* 6 months & *<* 12 months) and fast-progressors (*<* 6 months). The distribution of predicted scores for each group shows that the model is consistent in assigning these scores to the patients regardless of the fold and that the predicted scores are proportional to the progression time (see Figure S3).

For the MSK NSCLC datasets (Samstein et al. [2019]), we have found that clinical transformer-based models performed significantly better (Figure 3) than regularized CoxPH, XGBoost, DeepSurv, Hu et al Default (with original hyperparameters as listed in the paper) and Hu et al Adj (hyperparameters adjusted to closely match clinical transformer’s). The results from shuffled input features and target clinical endpoints (mean c-index of 0.51 *±* 0.097 and 0.48 *±* 0.092 respectively) demonstrate that clinical transformer recognizes the negative impact on performance when underlying input-output relationships are significantly changed. Clinical transformer without transfer learning (mean c-index of 0.59 *±* 0.094) showed significantly worse performance compared to clinical transformer models with transfer learning (mean c-indexes of 0.61 *±* 0.099 and 0.61 *±* 0.082). We noted insignificant reduction in performance for the clinical transformer with snapshot compared to the one without (P = 0.56). However, the model without the snapshot (TL) demonstrated a bias towards those variables that are related to the other cancer types when applied to NSCLC. This problem was mitigated by introducing a “snapshot” of NSCLC patients to the initial transfer learning phase (see Figure 1 for more details). We chose clinical transformer (Transfer Learning with snapshot) as our main model and regularized multivariate CoxPH as the baseline model. The hazard ratios from the main model were significantly better (that is, lower values) (Figure 4) than those from CoxPH model (N=59 patients, Mann-Whitney P<0.0006). We also observed that the predicted scores from clinical transformer showed better patient stratification compared to scores from the CoxPH model (Figure 4B and C). Clinical transformer model was also consistent in assigning scores to the 4 survival groups (Figure S3).

**Figure 3:**
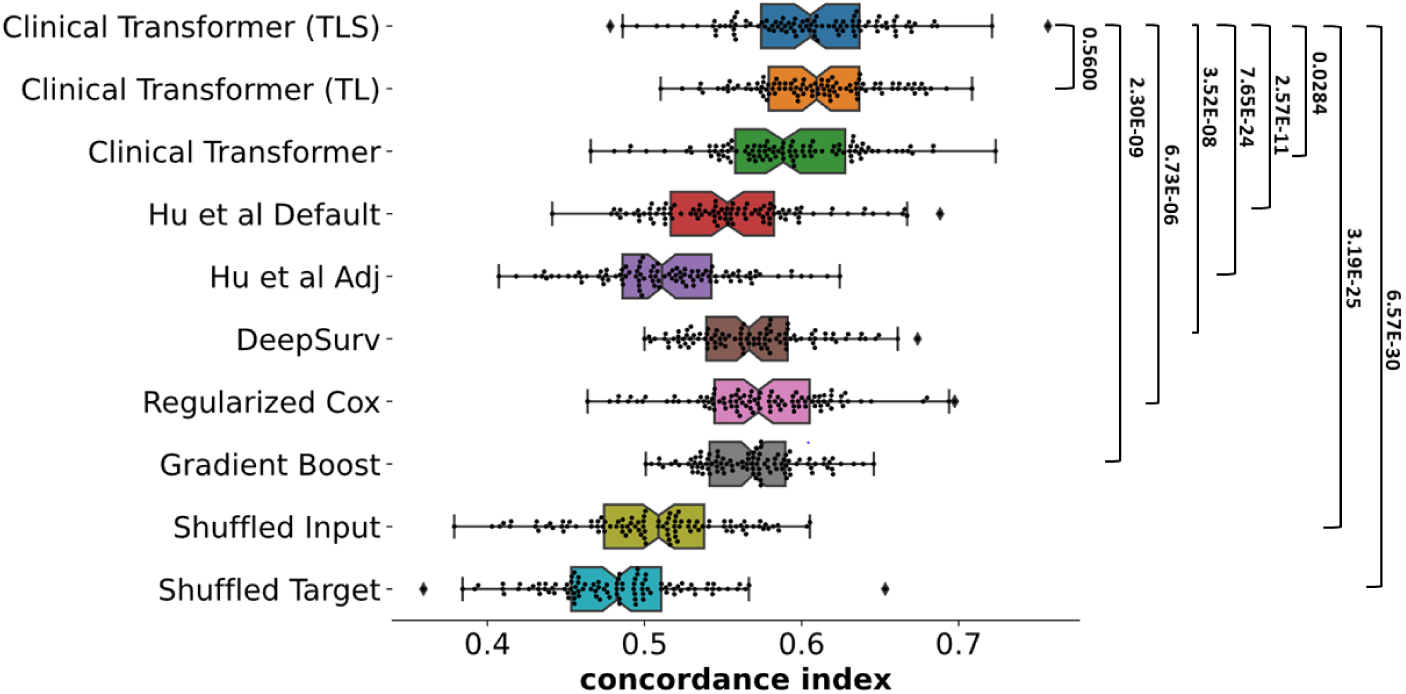
Concordance index comparison between different models from 100 different training-test random splits. The c-index distribution from transfer learning clinical transformer with snapshot (TLS) is compared to the c-index from the other models using Mann Whitney U two-sided test.

**Figure 4:**
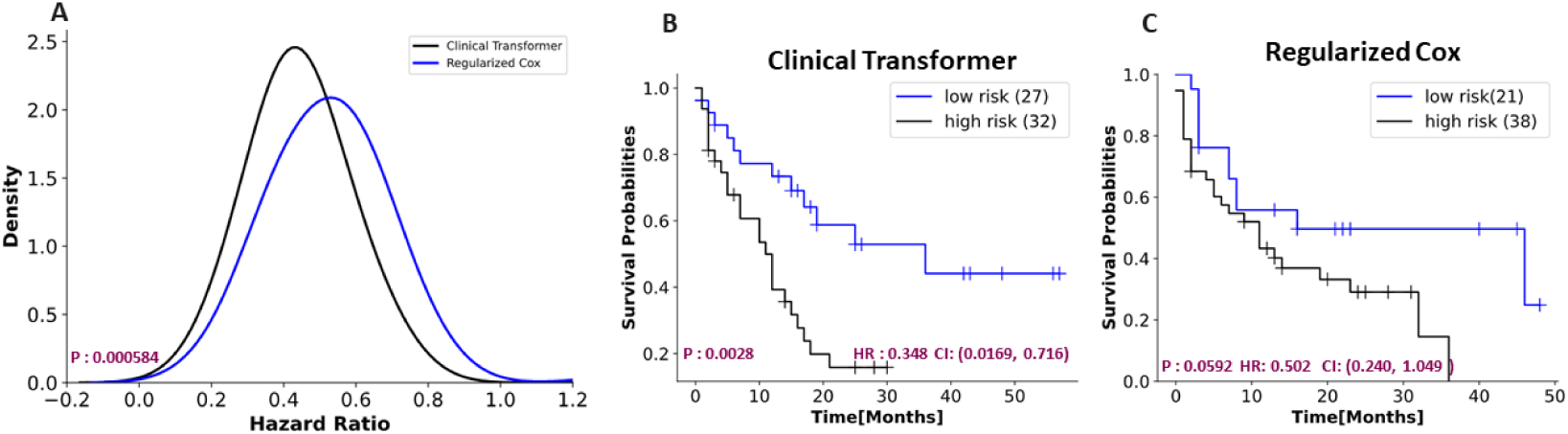
Evaluation of the predicted scores from clinical transformer and regularized cox model. **A**, Hazard ratios (HR) of predicted scores from the two models were computed for all the 100 training-test random splits. The HRs were computed using the optimal cut-off that maximizes the separation between high risk and low risk patients. Hazard ratios from clinical transformer were significantly lower than those from cox model (N=59 patients, Mann-Whitney P< 0.0006). **B** *−* **C**, Predicted scores from clinical transformer showed better patient stratification compared to the scores from cox model. While the two models show that patients with low-risk scores tend to have longer survival, clinical transformer shows that this group has significantly longer survival (HR=0.348, 95% CI:0.0169-0.716, log-rank P=0.0028). The two Kaplan Meier plots were generated from a single randomly selected fold (see Figure S1 and S2 for more examples)

### 4.2 Transfer learning based on cross-domain data improves the performance of domain-specific transformers

We sought to test if transfer learning could improve the prediction of overall survival that was based only on domain-specific data set. To train a cross-domain clinical transformer (Figure 2B) that is independent of the data set utilized to train the domain-specific transformer, we used data from ten different cancer types (N = 1266). The weights from this model were then used to initialize a NSCLC cancer-specific transformer. (Figure 3) demonstrates that the clinical transformer with the transfer learning is significantly better than the clinical transformer without transfer learning (“clinical transformer with transfer learning” with or without a snapshot vs. “clinical transformer – default”, with a p-val < 0.0035 and p-val < 0.03 respectively (Table S2)). These findings indicate the potential advantage of transfer learning to improve the ability to use the clinical transformers in clinical settings where a relatively small data set is available for analysis.

### 4.3 Interpretation of transformer predictions derived from attention weights

In contrast to linear models (e.g. CoxPH), where the importance of independent variables can only be explicitly derived, in models such as deep neural networks, that can be achieved only implicitly. Here, we demonstrate how the attention weights in the clinical transformer could be used to measure the strength of association of the variables with other variables and their effect on the prediction of the clinical outcome. To summarize the attention scores, we propose the Variable Interaction Score (VIS) that considers the interactions among the input features and the association with survival.

The *V IS* for a given population *P* and a variable *f* is defined as the sum of all the top attention pairs between input features *F* normalized by the number of patients (*P*) across the attention heads *H* for a given layer *L*:

- Attention matrices in the layer *L* are aggregated by heads *H* and population *P* to obtain a single matrix of averaged attention per population (or a single patient when *P* = 1).
- Then top ten interactions scores from each input variable *f* are selected
- The *V IS* for variable *f* is the sum of these interaction scores.

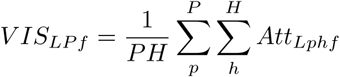

Since the *V IS* can be calculated at the single patient level, this metric can capture in addition to prevalent population of patients, a rare event – a small group of patients with a strong prognostic or predictive molecular or clinical characteristics.

### 4.4 Feature importance recapitulates current knowledge

To understand the extent to which our clinical transformer captures known cancer biology, we ordered the features used in the model by VIS (the measurement of importance described earlier in the text) and compared the top ones to known biology. (Figure 5) describes the importance of each variable in the model. As expected, the most important gene according to VIS for response to immunotherapy is TP53. TP53 is a key member in the cell – it encodes the p53 protein which increases under cell stress and is involved in multiple biological pathways including senescence, DNA repair and apoptosis (Aubrey et al. [2016]). As results of deleterious mutations in TP53, cells in the body accumulate DNA mutations. This abnormal process increases the prevalence of neoantigen production which may stimulates the immune response in the tumor microenvironment and thus introduce an advantage for immune-therapy. It has been previously reported by Dong et al. [2017], Lin et al. [2020], that TP53 mutations can be used as a prognostic biomarkers for improved outcome in immune checkpoint inhibitors. Notably, the other top hits that came up based on the VIS metric have been proposed as biomarkers for immunotherapy for example TMB (Greillier et al. [2018]), MGA (Sun et al. [2021]) and KEAP1 (Papillon-Cavanagh et al. [2020]). Further, we examined whether the CoxPH and the clinical transformer models accorded similar levels of importance to the input features. Although the two models are different in terms of their underlying modelling assumptions and complexity level, we expected to see some level of agreement in variable rankings, especially in well-known biomarkers in Immunotherapy. A summary of the comparison results is shown in Figure S4. We found a reasonable concordance in the importance levels of the variables between the two models (Pearson correlation of 0.53). Notably, the prominent features such as TMB (Samstein et al. [2019]) and TP53 associated with response to treatment for NSCLC exhibited a high rank in both models (Lin et al. [2020]).

**Figure 5:**
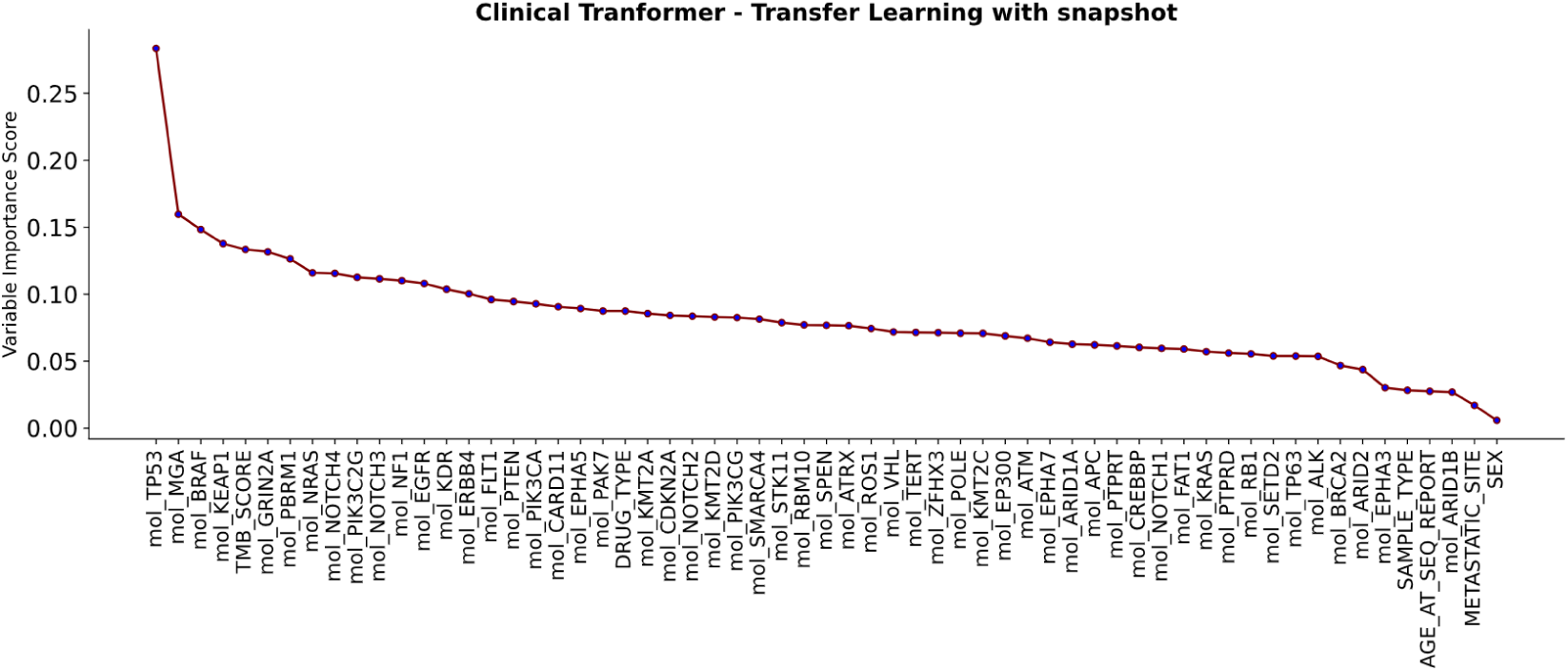
Feature importance ranked by the median absolute magnitude of Variable Interaction Score (VIS). Clinical transformer ranks both TMB and TP53 among the most important variables, recapitulating known findings on strong association between these features and survival outcomes for NSCLC patients. We denoted some variables with truncated name where necessary, otherwise, we kept the original notation as found in the datasets

### 4.5 Attention weights interpretation reveals key biological insights

Finally, we sought to test the possible connection of the attention score with the underlying tumor biology considering the survival time of the patients. A common practice in biology is to explore the patterns arising from data derived from biological experiments in different contexts (the overall survival time in this work) (Brunet et al. [2004]). For example, mRNA gene expression measured across many samples is used to calculate the correlation matrix between all gene pairs. This matrix is then used downstream to explore co-expression of genes through cluster analysis and can be associated with the conditions of the performed experiments (that is, the context) (Langfelder and Horvath [2008], Van Dam et al. [2018]).

A primary interest in clinical trials is to identify biomarkers for a given treatment that can predict fast-progressor and supper-responders; the former since those patients may benefit from a different class of anticancer drugs; and the latter because those are the true intent-to-treat population that would most benefit from immunotherapy. As done earlier in this work, we split the patient population into four ‘survival’ groups. Since these four groups respond differently to the treatment, biological functions that are known to be strongly associated with cancer progression may contribute to these sub-populations in different magnitude. Therefore, we grouped the genomic features (that is, the genes) in our data based on their known function to two different key functionalities that are strongly related to cancer progression; onco-suppressors genes (Repana et al. [2019]) and growth pathways genes (Reimand et al. [2019]). Furthermore, since the genes measured in our data sets, were pre-selected based on their strong association with cancer (see Figure 1 for further details), we included a set of all genes as another group in our analysis (Figures S8A). Lastly, we grouped three key clinical features together to assess their differential interactions across the four patients’ sub-populations. As expected, these four feature groups demonstrate monotonic decrease (Figure 6A and Figures S8A,S8B) or increase (Figure 6B) as a function of patient survival. We are currently exploring the incorporation of VIS into pathway interaction networks and other common practices in systems biology.

**Figure 6:**
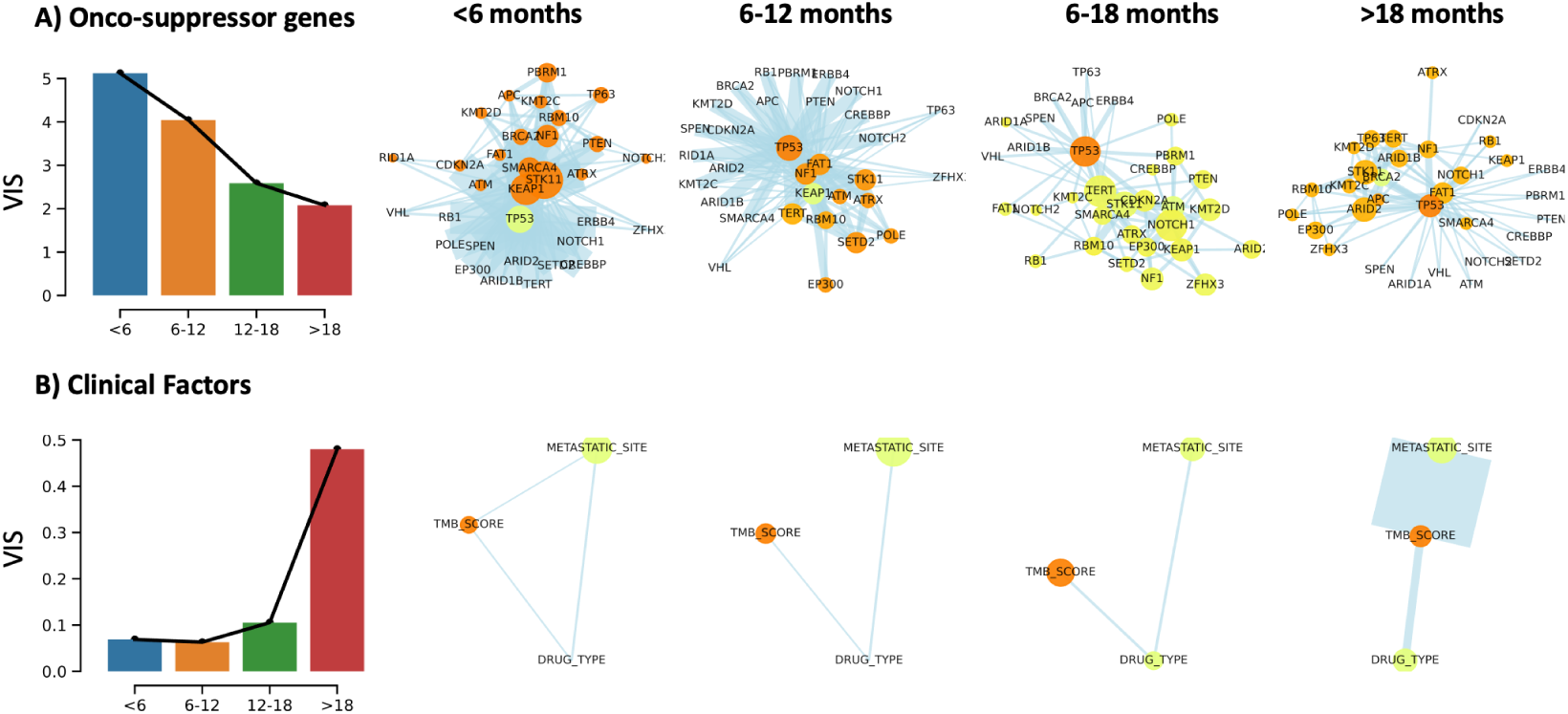
Attention weights reveal biological insights. Test population is divided into four intervals according to their survival time as indicated in the main text. Self-attention score (attention that is given to the same variable, or the diagonal of the attention matrix) is represented by the color of the nodes (yellow: low attention, orange: high attention). Edges represent variable interactions and their thickness describes the level of VIS (described in the main text). Sizes of the node represent the relative value of the input variable (e.g., number of mutations, TMB score).

## 5 Summary

In this work we have introduced a utilization of the attention mechanism for clinical applications in oncology. Cancer is a complex disease with wide variations across patients and many random events which accumulate in the lifetime of the disease. In addition, each patient has a different profile, at the molecular level (e.g., germline mutations) and physical (e.g., immune system, body mass and more). Here, we have demonstrated the ability of the clinical transformer to capture the semantics in patients’ profiles in the context of their survival. We have shown the superiority of this approach over other methods and the added value of using the interaction scores between features to bring system level insights. Furthermore, using a variant of transfer learning in our model, an option that is not feasible in static ML methods like coxPH and others, we have demonstrated a significant improvement over the baseline clinical transformer. Hence, enabling better modelling of small cohorts in clinical studies.

### 5.1 Limitations

As it is well established, the dot-product attention operation inside the transformer uses *O*(*n*^2^) time and space making it computational expensive for long input feature spaces. Therefore, the main limitation of the clinical transformer in its current implementation is the input feature dimension with a hard cap of 300 features. However, alternative strategies such as the low rank matrix attention proposed in the LinFormer Wang et al. [2020] can be used to reduce the time and space complexity to O(n) in the clinical transformer. Moreover, cancer treatment outcomes can be affected by various factors including patient fitness, tumor stage, pharmacokinetics, pharmacodynamics, tumor microenvironment and tumor molecular characteristic, among others. As in many clinical studies, including the studies analyzed here, there were only a sub-set of clinical and mutational features available. We recognize that these do not cover the complete underlying system complexity, which in many areas of biology is the case. Lastly, an important aspect in clinical trials is to find predictive biomarkers - a function that changes the hazard rate depending on the treatment. Discovering predictive biomarkers requires studies with at least two arms (one of which is a control group). In this work, we have analyzed data comprising a single family of treatments that is not from a randomized study. Thus, our findings here could indicate a prognostic value rather then predictive.

### 5.2 Broader impact

The personalization of clinical treatment is increasingly becoming critical towards expanding the delivery of optimum healthcare and enhancing the quality of life for individuals, particularly those with severe illnesses that require pro-active diagnosis and management. The ability to automate survival prognosis at an early stage at a personalized level has been a key goal in precision medicine because the early identification of potential trajectories of such patients and factors influencing their outcomes has vital ramifications towards optimal treatment planning. Algorithmic automation in this regard lowers the barriers to entry for such precision medicine systems and potentially extends to point-of-care applications in the developing world as well. A potential source of risk towards widespread deployment may stem from the demographic compositions in source datasets, and future translational studies would aim at curating geographically diverse datasets towards studying such effects comprehensively and mitigating any unwanted biases to that end.

## Data Availability

All data produced in the present study are available upon reasonable request to the authors

https://www.cbioportal.org/study/summary?id=msk_impact_2017

## 5.3 Availability

Code will be available upon request to the authors

## Notes

### Competing Interest Statement

The authors have declared no competing interest.

### Funding Statement

This computational study was funded by AstraZeneca - Early Computational Oncology and the graduate program

