## Supplementary File for "Explainable Transformer-Based Neural Network for the Prediction of Survival Outcomes in Non-Small Cell Lung Cancer (NSCLC)"

**Text related to Figure 1:**

In current clinical practice, the majority of precision oncology platforms use next-generation sequencing of targeted gene panels (that is, a subset of cancer related genes that is pre-decided to be sequenced out of approximately 20,000 genes in the human genome). The data that was used in this study (download from https://www.cbioportal.org/study/summary?id=tmb_mskcc_2018) is based on the clinical care at Memorial Sloan Kettering Cancer Center (MSK). Patients undergo genomic profiling with the Food & Drug Administration (FDA)-authorized Integrated Mutation Profiling of Actionable Cancer Targets (MSK-IMPACT) assay12. This test is performed in a Clinical Laboratory Improvement Amendments (CLIA)-certified laboratory environment and identifies somatic exonic mutations in a predefined subset of 468 cancer-related genes, by using both tumor-derived and matched germline normal DNA. See Samstein et al. [2019] for more details.

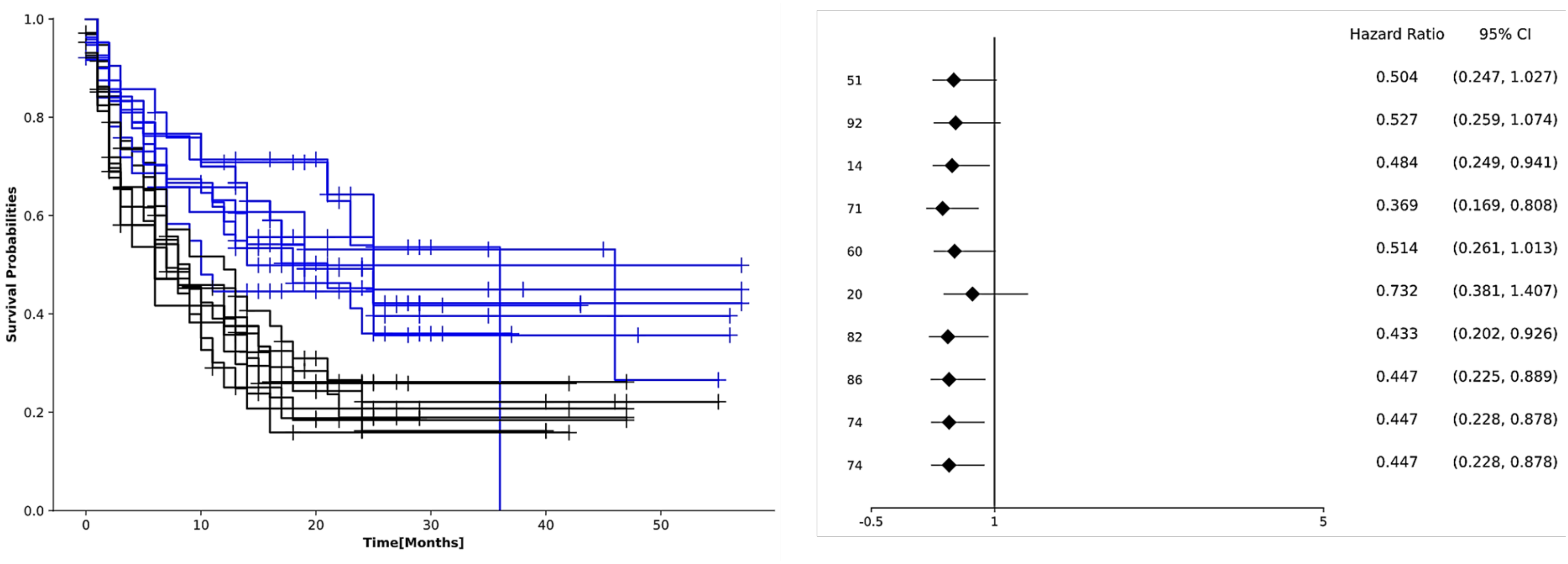


**Figure S1**. Patient stratification based on predicted scores from **clinical transformer**. The Kaplan Meier plot shows stratification of patients to low (black) and high (blue) scores groups in 10 randomly selects folds out of the 100. The plots are generated by searching for the optimal cut-off of the predicted scores so as to maximize the separation between the two groups. The hazard ratios and correspond 95% confidence intervals comparing low vs high score groups are computed. A confidence interval that does not cross the vertical line at 1 indicates that the corresponding two Kaplan Meier curves are significantly different. We observe better patient stratification when using predicted scores from clinical transformer compared to those from regularized cox model (Figure S2).

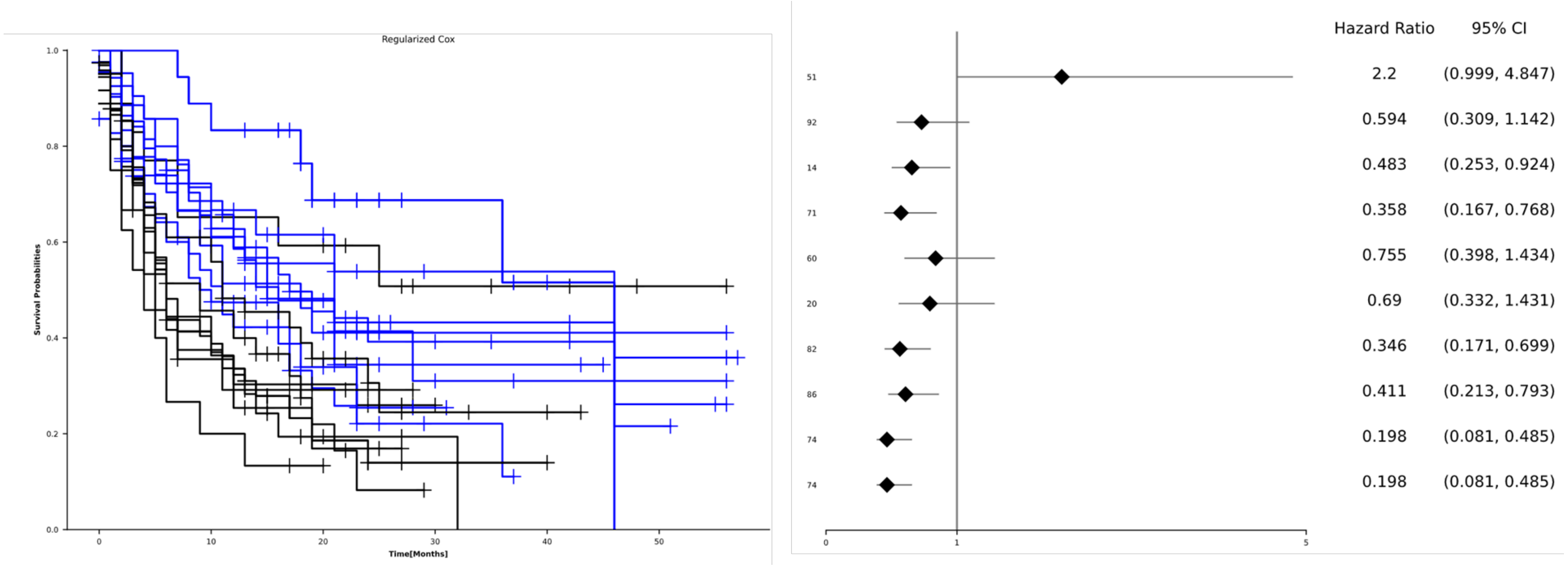


**Figure S2**. Patient stratification based on predicted risk scores from **regularized Cox** model. The Kaplan Meier plot shows stratification of patients to high (black) and low (blue) risk groups in 10 randomly selects folds out of the 100. The plots are generated by searching for the optimal cut-off of the risk scores so as to maximize the separation between the two groups. The hazard ratios and correspond 95% confidence intervals comparing low vs high risk are computed.

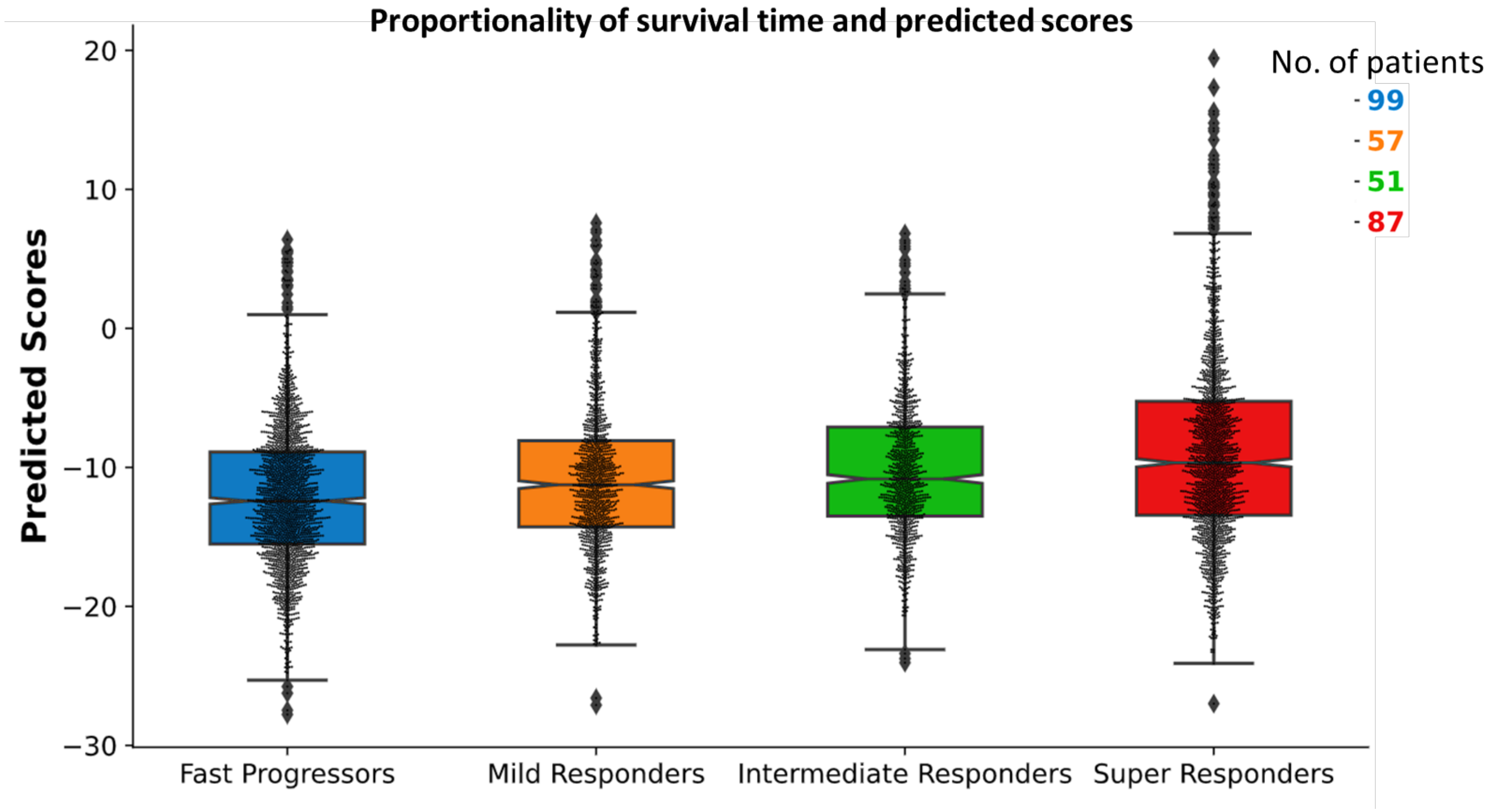

Proportionality of survival time and predicted scores
No. of patients
99
57
51
87
Predicted Scores
20
10
0
−10
−20
−30
Fast Progressors
Mild Responders
Intermediate Responders
Super Responders

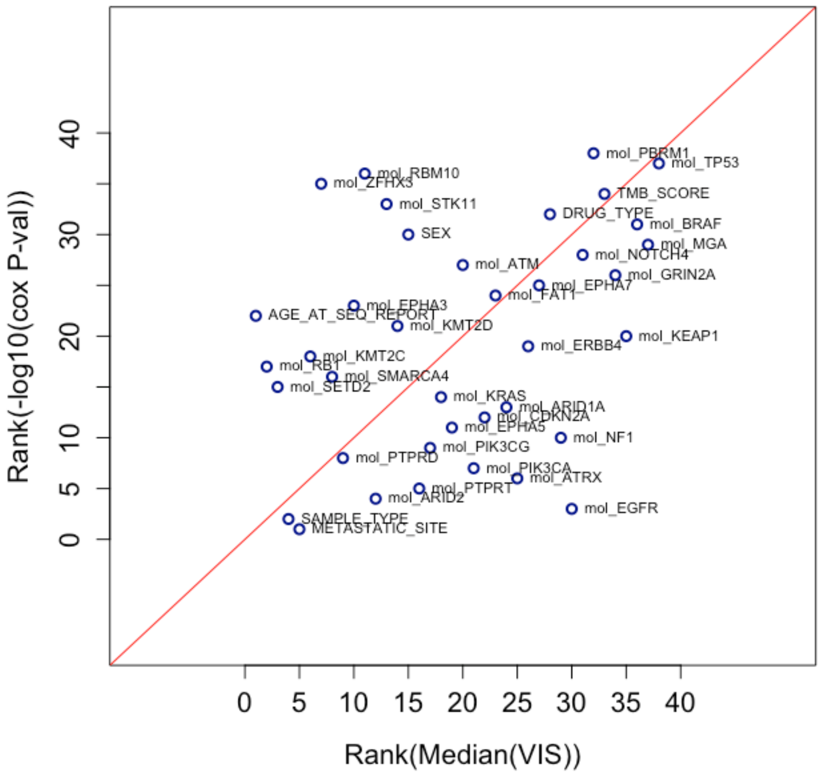


**Figure S4**. Comparison between the results of standard practice in survival analysis and the clinical transformer. Y-axis represents the ranks ordered by feature importance of the Cox proportional hazards model with L1 regularization. X-axis represents the ranks ordered by the feature importance derived from the VIS score of the clinical transformers. See the section “Interpretation of transformer predictions derived from attention weights” for further details on how the VIS is calculated and how the bootstrapping was performed. The Pearson correlation coefficient between the mean of the rank of -log10 p-values derived from the cox PH model and the rank of the median of the VIS score is 0.53 with a p-value < 0.0006. Only 38 features that were shared between the two models were used in this analysis. The non-overlapping variables were either constant (same value for all patients) or too sparse (<5% with non-zero entry) for training-test splits.

| **Model** | **Mean c-index ± 2 standard deviations** |
|---|---|
| Clinical Transformer (Transfer Learning with snapshot) | 0.61 ± 0.099 |
| Clinical Transformer (Transfer Learning without snapshot) | 0.61 ± 0.082 |
| Clinical Transformer - Default | 0.59 ± 0.094 |
| Hu et al Default | 0.56 ± 0.096 |
| Hu et al Adj | 0.51 ± 0.089 |
| DeepSurv | 0.57 ± 0.078 |
| Regularized Cox | 0.57 ± 0.091 |
| Gradient Boost | 0.57 ± 0.064 |
| Shuffled Input | 0.51 ± 0.097 |
| Shuffled Target | 0.48 ± 0.092 |

**Table S1.** Test set C -indexes for the different models. The dataset is split into 80% training and 20 test sets. Each model is trained on the 80% and its performance evaluated in terms of concordance index using the test set. To assess model's stability, the process is repeated on different splits 100 times. The corresponding c-indexes for the models are averaged and standard deviation computed.

**Mann Whitney U test on c-index between different models**

| | Clinical Transformer (Transfer Learning with snapshot) | Clinical Transformer (Transfer Learning without snapshot) | Clinical Transformer - Default | Hu et al Default | Hu et al Adj | DeepSurv | Regularized Cox | Gradient Boost |
|---|---|---|---|---|---|---|---|---|
| Clinical Transformer (Transfer Learning with snapshot) | | 0.5601 | 0.0284 | 2.57E-11 | 7.65E-24 | 3.52E-08 | 6.73E-06 | 2.30E-09 |
| Clinical Transformer (Transfer Learning without snapshot) | | | 0.0035 | 4.07E-14 | 2.47E-27 | 8.53E-11 | 7.09E-08 | 1.52E-12 |
| Clinical Transformer - Default | | | | 3.17E-07 | 9.21E-21 | 0.00024 | 0.010632 | 5.75E-05 |

**Table S2**. Mann Whitney U test on testing set c-indexes. P-values in the intersection of the respective models
